# Association between prostate cancer and myocardial infarction management and post-infarction outcomes: A Norwegian registry study

**DOI:** 10.1101/2024.11.04.24316728

**Authors:** Rachel B Forster, Camilla Kjellstadli, Rupali Akerkar, Gerhard E Sulo, Tor Åge Myklebust, Øystein Karlstad, Tone Bjørge, Kaare H Bønaa, Ester Kringeland, Rune Kvåle

## Abstract

**Background and aims:** Prostate cancer (PCa) is the most frequently diagnosed cancer in men in Norway and as survival rates improve cardiovascular disease (CVD) has emerged as a primary cause of morbidity and mortality, including acute myocardial infarction (AMI). Cancer and CVD share some important risk factors and PCa treatment may increase the risk of CVD. The aim of this study was to compare rates of invasive management, in-hospital complications, major adverse cardiovascular events (MACE), re-infarction and death, as well as prescription of guideline recommended secondary pharmacological prevention after an AMI between PCa patients and the general male AMI population.

**Methods:** Data included nation-wide registry data to identify all males 40-85 years in Norway who had their first AMI during 2013-2019. We compared outcomes after AMI between those diagnosed with localized PCa between 2004-2019 and the general AMI population using logistic and cause-specific Cox regression.

**Results:** 34,362 AMI patients were included, of whom 1405 (4.1%) had PCa. No differences were observed in invasive management or secondary medical treatment post-AMI between PCa patients and non-cancer patients. While PCa patients had a lower risk of overall complications (OR 0.77; 0.64-0.92), they experienced an increased risk of serious bleeding (OR 1.66; 1.08-2.44) and no difference in MACE or re-infarction events. PCa patients had better 1-year survival (HR 0.82; 0.69-0.98).

**Conclusions:** There was no evidence of reduced quality of AMI care for PCa patients in Norway. These findings support treatment of AMI as usual for localized PCa patients, but with attention to increased bleeding risk.

**Key learning points:** *What is known:*

- Evidence from studies evaluating quality of care and outcomes of <u>cancer patients after an acute myocardial infarction</u> have found that those with cancer are more likely to receive poorer quality of care and have worse outcomes, compared to non-cancer patients.
- Prostate cancer is one of the most frequently diagnosed cancers in men, and as survival rates improve, cardiovascular disease has emerged as a primary cause of morbidity and mortality, including acute myocardial infarction.

*What the study adds:*

- There was no evidence that non-metastatic prostate cancer patients receive reduced quality of care when they experienced an acute myocardial infarction or that they were at higher risk of adverse outcomes in the following year.
- Overall, risk of in-hospital complications was lower in prostate cancer patients, except for serious bleeding, which was more likely.
- Non-metastatic prostate cancer patients should receive usual treatment for a myocardial infarction but with individualized consideration of their higher risk of bleeding.

*Non-standard abbreviations and acronyms:* ADT = androgen deprivation therapy; CCI = Charlson comorbidity index; NorPD = Norwegian Prescription Database; NORMI = Norwegian Myocardial Infarction Quality Registry PCa = prostate cancer

## Introduction

Prostate cancer (PCa) is one of the most common cancers globally, with particularly high incidence rates in Northern Europe ^(1, 2)^. Between 2012 and 2016 Norway had the second highest age-standardized incidence rate in Europe ^(2)^. As survival rates improve due to advancements in cancer detection and treatment, more cancer survivors are at risk of developing and dying from cardiovascular disease (CVD), which is the leading non-cancer cause of death in men with PCa ^(3, 4)^. At the time of PCa diagnosis, patients commonly have coexisting CVD, which share risk factors, including age, inflammation and lifestyle factors ^(5–7)^. In addition to the improved survival rate after PCa, other factors such as the cancer itself and treatment with androgen deprivation therapy (ADT) and radiation therapy, may increase the risk of developing CVD, including acute myocardial infarction (AMI) ^(7–10)^.

Historically, trials for the management of AMI have largely been restricted to patients without active cancer, despite the growing overlapping population of patients with cancer and AMI ^(11, 12)^. In recent studies, cancer patients have been reported to be less likely to receive invasive management for AMI, have worse outcomes after AMI, and less likely to receive secondary pharmacological prevention than the general population ^(11, 13–16)^. Worse AMI treatment in cancer patients is likely influenced by the increased risk of bleeding from AMI treatment ^(6)^. However, these studies on AMI management in cancer patients have been conducted in populations with mixed cancer types.

Prior European Society of Cardiology (ESC) guidelines for non-ST-elevated AMI (NSTEMI) gave cancer as a reason to withhold invasive management, due to increased risk of bleeding, without further guidelines for treatment escalation, while the ST-elevated AMI (STEMI) guidelines did not mention cancer at all ^(17, 18)^. In contrast, the newest ESC guidelines on management of acute coronary syndromes and on cardio-oncology reflect a better understanding of AMI in cancer patients and recommend an invasive strategy for cancer patients with expected survival beyond six months ^(19, 20)^. There remains a gap in cancer specific knowledge in the event of an AMI to further inform these developing guidelines.

To address this knowledge gap, this study aims to compare rates of invasive management, in-hospital complications including serious bleeding, major adverse cardiovascular events (MACE), re-infarction and death, as well as rates of guideline recommended secondary pharmacological prevention after an AMI, between non-metastatic PCa patients and men without a history of cancer.

## Methods

### Study Design and Population

This study was a nationwide, registry-based, longitudinal cohort study, and included all males in Norway, aged 40 to 85 years, with an AMI from 2013 to 2019. People were excluded if they had suffered an AMI prior to this period or if they were diagnosed with any other type of cancer than PCa or non-melanoma skin cancer from 1953 to entry in the study. Individuals diagnosed with primary distant metastatic PCa were also excluded. The primary exposure of interest was the diagnosis of PCa from 2004 to 2019, and prior to the first occurrence of AMI. The study was approved by the Regional Committee for Medical Research South-East in Norway (130363).

### Data sources

Data for this study were obtained from the Cancer Registry of Norway specific to PCa cases (International Classification of Diseases-10 (ICD-10) C61) as well as other types of cancer in the whole male population, linked using the personal identification number allocated to all residents in Norway. Additional clinically relevant information specific to PCa was obtained from the Norwegian Prostate Cancer Registry. Data on diagnosis and treatment of AMI (ICD-10 I21 and I22) and other CVDs were obtained from the Norwegian Cardiovascular Disease Registry, with detailed event information from the Norwegian Myocardial Infarction Quality Registry (NORMI). Prescription medications were obtained from the Norwegian Prescription Database (NorPD). Data on education was obtained from Statistics Norway. Additional health information to estimate comorbidity came from the Norwegian Patient Registry, and cause of death came from the Norwegian Cause of Death Registry.

### Outcome definitions

Using data from NORMI, in-hospital complications that occurred during hospitalization for the index AMI included cardiogenic shock, re-infarction, AV-block grade II-III, mechanical complications/rupture, heart failure and serious bleeding and were included in the analyses as a composite endpoint. As bleeding is a major concern for cancer patients, we also evaluated this outcome separately. Serious bleeding was defined as: any bleeding with fatal outcome; or any intracranial, intraspinal, retroperitoneal, intraocular, intra-articular, or pericardial bleeding causing clinical symptoms; or any bleeding associated with a fall in Hgb > 5 g/dl; or requiring transfusion of more than 1 unit of blood.

Invasive AMI management was defined as invasive examination (invasive coronary angiography), and invasive intervention (percutaneous coronary intervention (PCI) or coronary artery bypass graft (CABG) surgery).

Secondary pharmacological prevention included dual antiplatelet therapy (DAPT) of aspirin plus a P2Y12 inhibitor, lipid-lowering medications, angiotensin converting enzyme inhibitors or angiotensin receptor blockers (ACEi/ARB) and betablockers, prescribed at discharge or within 30 days of the AMI. Patients that died during hospitalization for AMI were excluded from this outcome.

Major adverse cardiovascular events (MACE) were derived as a composite of non-fatal stroke (ICD-10 I61, I63 and I64), non-fatal AMI (ICD-10 I21 and I22) and CVD death (underlying cause of death ICD-10 I00-I99). Re-infarction was defined as any AMI event, fatal or non-fatal, after the index AMI. All-cause mortality was defined as death from any cause after the index AMI event.

### Variables

Data on the patient’s age and year of AMI admission, as well as history of smoking, stroke, hypertension treatment, chronic heart failure, cardiac procedures, and diabetes mellitus were collected from NORMI at the time of the index AMI. Prior stroke, chronic heart failure, and cardiac procedures were grouped together as a composite, binary ‘prior CVD’ variable indicating if the patient had a history of any of these conditions or procedures.

Charlson comorbidity index (CCI) was calculated using data from Norwegian Patient Registry in the 2 years prior to the index AMI ^(21)^. The score excluded cancer and myocardial infarction from the calculation and was categorized as 0, 1, 2, 3 and 4+. Highest education reported in the 3 years prior to the index AMI event was identified from Statistics Norway and reported as primary, secondary or tertiary education.

Age and year of PCa diagnosis, Gleason score, clinical staging of the primary tumour (cTNM), prostate-specific antigen (PSA) at diagnosis and primary treatment information were collected from the Norwegian Prostate Cancer Registry. Information on hormonal PCa treatment was obtained from NorPD.

Primary PCa treatment was defined as active surveillance, radical prostatectomy, definitive radiotherapy (RT) (target dose of ≥74 Gy in 2 Gy units) and ADT, starting within 6 months after diagnosis. For combination ADT and RT, RT could begin max 6 months after ADT start to be considered primary PCa treatment. To fulfil the criteria for active surveillance the patient had to be clinical stage ≤T2a, Gleason ≤7a, could not have distant metastatic disease, and have not received a prostatectomy, RT or ADT in the first 6 months after diagnosis.

We calculated risk groups for recurrence based on the European Association of Urology (EAU) guidelines, which are used in clinical practice in Norway ^(22, 23)^. Low-risk disease was defined as Gleason score ≤6 and PSA <10 ng/mL and cT1-2a; moderate-risk: Gleason score 7 or PSA 10–20 ng/mL or cT2b; and high-risk disease: Gleason Score >7 or PSA >20 ng/mL or cT2c; locally advanced disease: cT3–4 or cN+.

### Statistical analysis

General characteristics were summarized using median and interquartile range (IQR) or numbers with percentage. Data were presented stratified by PCa diagnosis status and then also within PCa patients, based on if they received primary ADT (p-ADT). To better understand differences in descriptive statistics between comparable age groups, general characteristics were presented by broad age groups (40-65 , 66-75 and 76-85 years) in **Supplemental 1**.

Outcomes including in-hospital complications, invasive management, and secondary prevention were analyzed using logistic regression, comparing PCa diagnosis to the general AMI population without PCa. Results were presented as odds ratios (OR) with 95% confidence intervals (CIs).

MACE, re-infarction, and all-cause mortality were visualized using cumulative incidence curves that include competing risks for MACE (death from non-CVD cause) and re-infarction (death from any cause). The cumulative incidence curves were stratified by age groups when comparing PCa to the general AMI population. Outcomes at 30 days and 1 year were analyzed using Cox regression, also accounting for competing risks, and results presented as cause-specific hazard ratios (HRs) with 95% CIs.

Additionally, for MACE, re-infarction and mortality outcomes, analyses were performed just in the PCa AMI population, comparing p-ADT treatment to no p-ADT treatment. These analyses were limited to the PCa population in the moderate-, high-risk and locally advanced EAU risk groups, and where outcomes occurred at least 6 months after PCa diagnosis to ensure treatment had started. Low-risk PCa patients were excluded as ADT is not recommended to this group.

Analyses were carried out in two stages, first adjusting for age at AMI (age-adjusted), and then including other covariates (fully-adjusted): age, smoking, prior CVD, hypertension treatment, diabetes, AMI year, education and CCI. Analyses for complications and invasive management also included ST-elevation, and the serious bleeding outcome additionally controlled for invasive treatment. The analyses carried out in the PCa population comparing primary treatment types additionally controlled for EAU risk group. The analyses of DAPT as secondary prevention additionally controlled for serious bleeding.

All analyses were performed using R version 4.3.0 ^(24)^.

## Results

### Population

A total of 34,362 males that experienced a first AMI between 2013 and 2019 and were registered in NORMI were included in the study, 1,405 (4.1%) of whom had a diagnosis of non-metastatic PCa prior to their AMI **(Supplemental 2)**. Median age of males with a PCa diagnosis was 73 (IQR 68.0-78.0) and 64 (IQR 55.5-72.5) years in the general AMI population **(Table 1)**.

**Table 1.** Population characteristics at time of AMI.

|  | **PCa**<br>**population**<br>**(n = 1405)** | **General AMI**<br>**population**<br>**(n = 32957)** | **p-ADT**<br>**(n = 361)** | **No p-ADT**<br>**(n = 678)** |
| --- | --- | --- | --- | --- |
| **Age- median**<br>**(IQR)** | 73 (68-78) | 64 (55.5-72.5) | 77 (72.5-81.5) | 73 (68-78) |
| **Age group** |  |  |  |  |
| **40-65** | 204 (14.5%) | 17589 (53.4%) | 24 (6.6%) | 90 (13.3%) |
| **66-75** | 650 (46.3%) | 9267 (28.1%) | 125 (34.6%) | 352 (51.9%) |
| **77-85** | 551 (39.2%) | 6101 (18.5%) | 212 (58.7%) | 236 (34.8%) |
| **ST elevation**<br>**status** |  |  |  |  |
| **Non-STEMI** | 941 (67.0%) | 19947 (60.5%) | 268 (74.2%) | 429 (63.3%) |
| **STEMI** | 406 (28.9%) | 11570 (35.1%) | 74 (20.5%) | 219 (32.3%) |
| **Died during**<br>**admission** | 67 (4.8%) | 1320 (4.0%) | 19 (5.3%) | 35 (5.2%) |
| **Prior diabetes** | 239 (17.0%) | 5481 (16.6%) | 70 (19.4%) | 113 (16.7%) |
| **Prior**<br>**hypertension**<br>**treatment** | 708 (50.4%) | 13056 (39.6%) | 192 (53.2%) | 330 (48.7%) |
| **Prior CVD** | 277 (19.7%) | 4950 (15.0%) | 82 (22.7%) | 130 (19.2%) |
| **Prior heart failure** | 32 (2.3%) | 747 (2.3%) | 15 (4.2%) | 8 (1.2%) |
| **Prior stroke** | 109 (7.8%) | 1820 (5.5%) | 40 (11.1%) | 48 (7.1%) |
| **CCI** |  |  |  |  |
| **0** | 728 (51.8%) | 19576 (59.4%) | 154 (42.7%) | 363 (53.5%) |
| **1** | 391 (27.5%) | 8277 (25.1%) | 105 (29.1%) | 201 (29.6%) |
| **2** | 169 (12.0%) | 3214 (9.8%) | 57 (15.8%) | 71 (10.5%) |
| **3** | 78 (5.6%) | 1187 (3.6%) | 29 (8.0%) | 29 (4.3%) |
| **4+** | 39 (2.8%) | 703 (2.1%) | 16 (4.4%) | 14 (2.1%) |
| **Current smoker** | 272 (19.4%) | 10802 (32.8%) | 64 (17.7%) | 131 (19.3%) |
| **Highest**<br>**education** |  |  |  |  |
| **Primary** | 365 (26.0%) | 9135 (27.7%) | 126 (34.9%) | 167 (24.6%) |
| **Secondary** | 732 (52.1%) | 16471 (50.0%) | 177 (49.0%) | 351 (51.8%) |
| **Tertiary** | 305 (21.7%) | 6916 (21.0%) | 58 (16.1%) | 159 (23.5%) |
| *Missing: ST elevation had 4.4% missing and current smoker had 7.0% missing data, all other variables had 1% or less missing data* |  |  |  |  |
| *AMI = acute myocardial infarction; CCI = Charlson comorbidity index; CVD = cardiovascular disease; IQR = interquartile range; p-ADT = primary androgen deprivation therapy; PCa = prostate cancer; STEMI = ST-elevated myocardial infarction* |  |  |  |  |

There were a total of 1,039 people in the PCa population specific analyses, after excluding low-risk and those with an AMI within 6 months after the PCa diagnosis. EAU moderate-risk comprised 47% (n = 489), high-risk 30% (n = 312) and locally advanced 23% (n = 238). Three-hundred and sixty-one (34.6%) received p-ADT treatment (**Table 2**).

**Table 2.** Prostate cancer characteristics.

|  | **PCa population**<br>**(n = 1405)** | **p-ADT**<br>**(n = 361)** | **No p-ADT**<br>**(n = 678)** |
| --- | --- | --- | --- |
| **Time from PCa diagnosis to AMI - median years (IQR)** | 5.3 (2.4-8.2) | 5.5 (2.8-8.3) | 5.6 (2.9-8.3) |
| **PSA- median (IQR)** | 9.5 (4.5-14.0) | 17.0 (11.5-28.5) | 10.1 (3.7-13.8) |
| **Gleason score** |  |  |  |
| **6** | 492 (35.0%) | 24 (6.6%) | 177 (26.1%) |
| **7a** | 382 (27.2%) | 107 (29.6%) | 255 (37.6%) |
| **7b** | 189 (13.5%) | 73 (20.2%) | 106 (15.6%) |
| **8** | 166 (11.8%) | 87 (24.1%) | 68 (10.1%) |
| **9** | 88 (6.3%) | 59 (16.3%) | 24 (3.5%) |
| **10** | 4 (0.3%) | 4 (1.1%) | 0 (0.0%) |
| **EAU risk group** |  |  |  |
| **Low** | 285 (20.3%) | - | - |
| **Moderate** | 514 (36.6%) | 90 (24.9%) | 399 (58.8%) |
| **High** | 327 (23.3%) | 132 (36.6%) | 180 (26.5%) |
| **Locally advanced** | 251 (17.9%) | 139 (38.5%) | 99 (14.6%) |
| **Primary treatment** |  |  |  |
| **ADT only** | 138 (9.8%) | 129 (35.7%) | - |
| **ADT + Radical prostatectomy** | 5 (0.4%) | 5 (1.4%) | - |
| **ADT + Radiotherapy** | 242 (17.2%) | 227 (62.9%) | - |
| **Active surveillance** | 248 (17.7%) | - | 44 (6.5%) |
| **Radical prostatectomy** | 377 (26.8%) | - | 297 (43.8%) |
| **Radiotherapy** | 118 (8.4%) | - | 103 (15.2%) |
| **No treatment** | 277 (19.7%) | - | 234 (34.5%) |
| **ADT Type (for primary treatment)** |  |  |  |
| **LHRH analoge** | 339 (24.1%) | 318 (88.1%) | - |
| **LHRH antagonist** | 6 (0.4%) | 5 (1.4%) | - |
| **Antiandrogen** | 356 (25.3%) | 334 (92.5%) | - |

*ADT = androgen deprivation therapy; EAU = European Association of Urology; LHRH = luteinizing hormone releasing hormone; IQR = interquartile range; p-ADT = primary ADT; PSA = prostate specific antigen*

### Invasive examination and treatment

In the PCa population, 1,160 (82.6%) had angiography and 1,023 (72.8%) received invasive intervention compared with 28,918 (87.7%) having angiography and 25,684 (77.9%) receiving invasive intervention in the general AM I population.

There were no associations between PCa and the general AMI population for receiving angiography (OR 1.10; 0.92-1.32) or invasive AMI treatment (OR 1.09; 0.94-1.26), after adjusting for potential confounders **(Table 3)**.

**Table 3.** Logistic regression results for outcomes invasive management and in-hospital complications comparing PCa to the general AMI population.

|  | **Outcome** | **Age-adjusted**<br>**OR (95% CI)** | **Fully-adjusted**<br>**OR (95% CI)** |
| --- | --- | --- | --- |
| **Invasive management** | Angiography | **1.22 (1.06-1.43)** | 1.10 (0.92-1.32) |
|  | Invasive treatment | **1.18 (1.04 - 1.33)** | 1.09 (0.94-1.26) |
| **In-hospital complications** | Composite complications | **0.78 (0.68-0.89)** | **0.77 (0.64-0.92)** |
|  | Serious bleeding | 1.39 (0.93-1.99) | **1.66 (1.08-2.44)** |
| *Fully adjusted models include age, smoking, prior CVD, hypertension treatment, diabetes, AMI year, education, CCI, ST-elevation and bleeding outcome additionally controlled for invasive treatment*<br>*CCI = Charlson comorbidity index; CI = confidence interval; CVD = cardiovascular disease; EAU = European Association of Urology; OR = odds ratio* |  |  |  |

### In-hospital complications

There were a total of 6,455 cases of composite complications (n = 262 with PCa) and 405 with serious bleeding (n = 30 with PCa).

For composite in-hospital complications, there was reduced odds among PCa patients compared to the general AMI population (OR 0.77; 0.64-0.92) **(Table 3)**. For serious bleeding, after controlling for potential confounders there was an increased odds of serious bleeding events in PCa patients compared to the general AMI population (OR 1.66; 1.08-2.44).

### MACE, re-infarction, and all-cause mortality at 30 days and 1 year

In the PCa population, the crude rate of MACE at 30 days was 7.3% (n = 102), re-infarction 1.4% (n = 20), and all-cause mortality 6.2% (n = 87), compared to the general AMI population with rates of 5.6% (n = 1,858), 1.1% (n = 349), and 5.1% (n = 1,694), respectively. At 1 year, the crude rates in the PCa population were 13.3% (n = 187) for MACE, 5.1% (n = 72) for re-infarction, and 12.0% (n = 168) for all-cause mortality compared with 10.2% (n = 3,343), 4.2% (n = 1370), and 9.1% (n = 3,000) in the general AMI population, respectively. Cause-specific cumulative incidence curves over 1 year after AMI are presented in **Figure 1, top panel,** stratified by PCa diagnosis and age-group.

**Figure 1.**
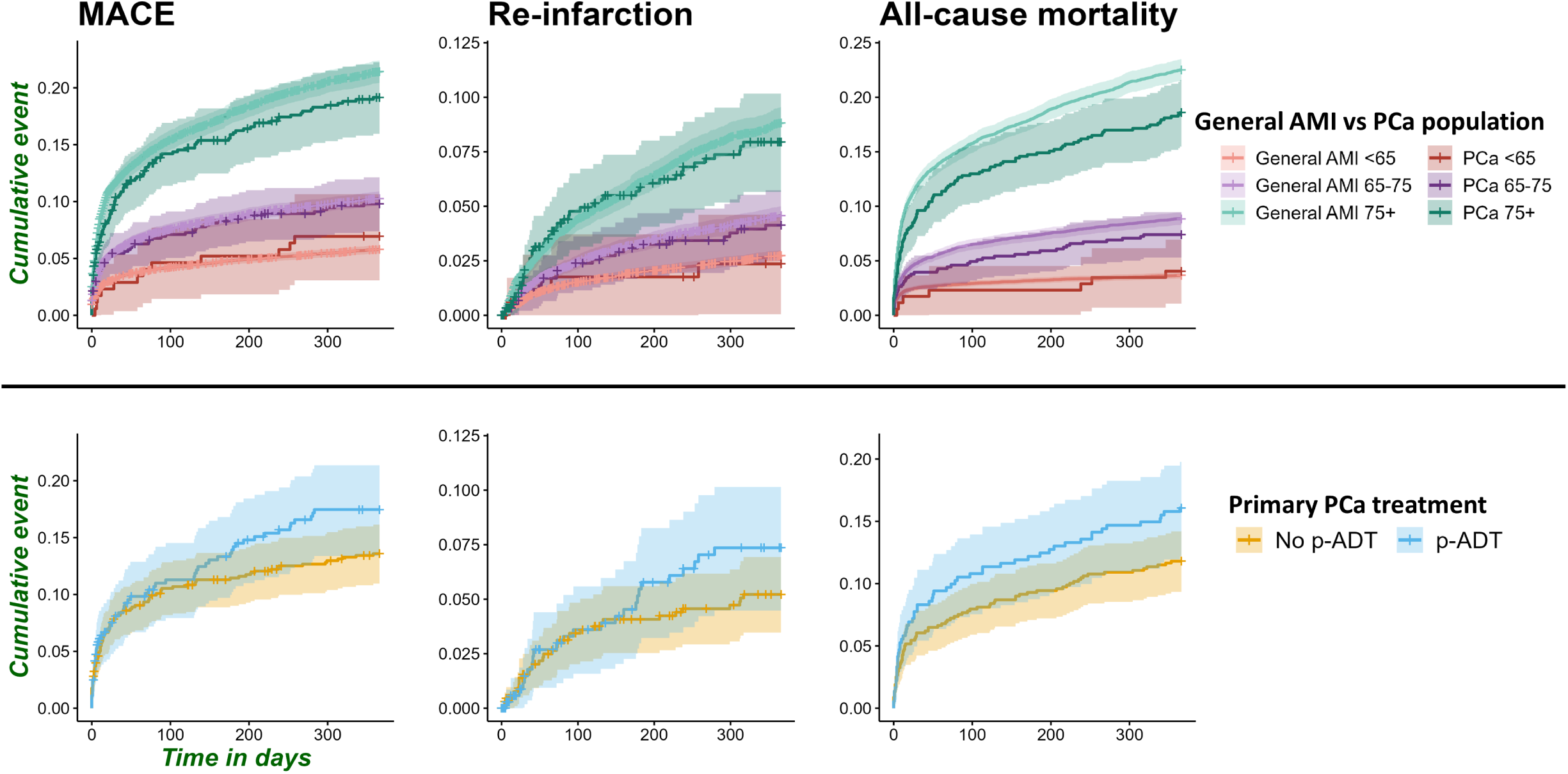
Cause-specific cumulative incidence curves up to 1 year after AMI for outcomes MACE, Re-infarction and all-cause mortality. **Top panel**: General AMI population vs PCa population, by age groups <65, 65-75 and 75+; presented by age groups as there were large differences in age between comparison groups; **Lower panel:** PCa only, comparing no primary ADT vs primary ADT; curves not provided in age groups as age was not largely different between the comparison groups AMI = acute myocardial infarction; MACE = major adverse cardiovascular event; p-ADT = primary androgen deprivation therapy; PCa = prostate cancer

There was no consistent pattern of association between PCa and MACE or re-infarction at 30 days and 1 year **(Table 4)**. The risk of all-cause mortality was reduced in the PCa population, with less precision at 30 days (HR 0.80; 0.62-1.04) compared to 1 year (HR 0.82; 0.69-0.98), after controlling for potential confounders. The unadjusted cumulative incidence curve for mortality shows this difference primarily in the older group, 75+ years **(Figure 1, top panel).**

**Table 4.** Cox regression results of outcomes MACE, re-infarction and all-cause mortality at 30 days and 1 year.

|  |  |  | Age-adjusted | Fully-adjusted |
| --- | --- | --- | --- | --- |
| Comparison | Time | Outcome | HR (95% CI) | HR (95% CI) |
| PCa v general AMI | 30 day | MACE | 0.89 (0.73-1.08) | 0.92 (0.73-1.16) |
|  |  | re-infarction | 1.02 (0.65-1.61) | 1.09 (0.69-1.75) |
|  |  | all-cause mortality | **0.77 (0.62-0.96)** | 0.80 (0.62-1.04) |
|  | 1 year | MACE | 0.90 (0.77-1.04) | 0.93 (0.79-1.10) |
|  |  | re-infarction | 0.89 (0.70-1.12) | 0.88 (0.68-1.14) |
|  |  | all-cause mortality | **0.80 (0.68-0.94)** | **0.82 (0.69-0.98)** |
| p-ADT v no p-ADT | 30 day | MACE | 0.84 (0.53-1.33) | 0.73 (0.43-1.27) |
|  |  | re-infarction | 0.75 (0.25-2.25) | - |
|  |  | all-cause mortality | 1.11 (0.69-1.81) | 0.88 (0.50-1.57) |
|  | 1 year | MACE | 1.06 (0.76-1.48) | 1.05 (0.72-1.52) |
|  |  | re-infarction | 1.08 (0.63-1.85) | - |
|  |  | all-cause mortality | 1.09 (0.77-1.54) | 0.91 (0.61-1.35) |
| Fully adjusted model includes age, smoking, prior CVD, hypertension treatment, diabetes, AMI year, education, CCI; ADT comparison additional controlled for EAU risk group |  |  |  |  |
| ADT = androgen deprivation therapy; AMI = acute myocardial infarction; CCI = Charlson comorbidity index; CI = confidence interval; CVD = cardiovascular disease; EAU = European Association of Urology; HR = hazard ratio; MACE = major adverse cardiovascular event; p-ADT = primary androgen deprivation treatment; PCa = prostate cancer |  |  |  |  |

In the PCa treatment analysis population, those that received p-ADT had crude rates of MACE at 30 days of 8.0% (n = 29), re-infarction in 1.1% (n = 5), and all-cause mortality of 8.3% (n = 30), compared to the no p-ADT population of 8.1% (n = 55), 1.5% (n = 10), and 6.0% (n = 41), respectively. After one-year, crude rates in the p-ADT group were 13.3% (n = 61) for MACE, 4.8% (n = 33) for re-infarction and 12.0% (n = 58) for all-cause mortality vs. 16.9% (n = 91), 6.1% (n = 24) and 16.6% (n = 80) in the no p-ADT group, respectively. Due to low numbers of re-infarction, only age-adjusted Cox models were carried out for this outcome. Among PCa patients, p-ADT did not have a significant impact on rates of MACE, re-infarction, or all-cause mortality at 30 days or 1 year **(Figure 1, lower panel; Table 4)**.

### Secondary pharmacological prevention

In the PCa population, DAPT was prescribed to 1,104 patients (82.5%), lipid-lowering treatment to 1,238 (92.5%), ACEi/ARB to 814 (60.8%), and beta-blockers to 957 patients (71.5%). In the general AMI population, DAPT, lipid-lowering treatment, ACEi/ARB, and beta-blockers were prescribed to 27,067 (85.6%), 29,600 (93.6%), 18,258 (57.7%), and 23,715 (75.0%) patients, respectively.

In age-adjusted logistic regression models, PCa patients were more likely to receive DAPT or lipid-lowering medications after an AMI (OR 1.23, 1.06-1.43 and 1.34; 1.09-1.66, respectively) and less likely to receive betablockers (OR 0.83; 0.73-0.94) (**Figure 2**). After controlling for potential confounding variables, the increased rate of lipid-lowering prescription in the PCa population remained (OR 1.27; 1.01-1.62) as did the reduced rate of betablocker prescription (OR 0.82; 0.72-0.93). There was no longer an association with DAPT and PCa status and no evidence of differences in prescriptions in ACEi/ARBs in any model.

**Figure 2.**
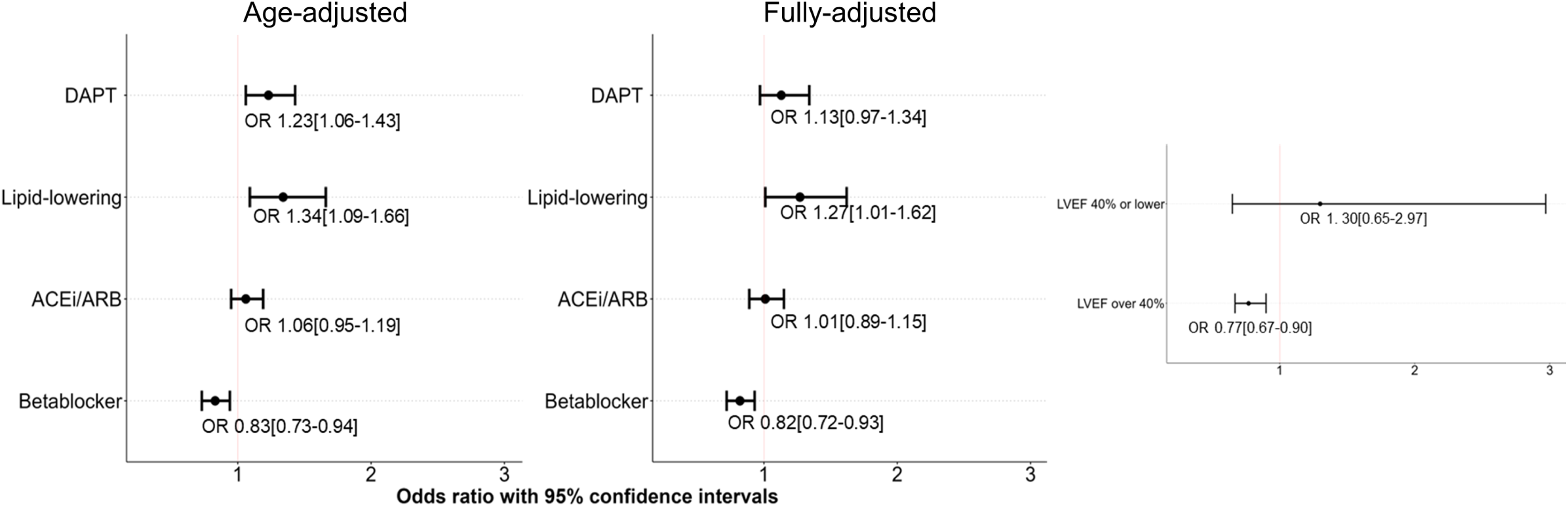
Logistic regression results for secondary prevention after AMI comparing PCa to general AMI population. **Left panel**: Age-adjusted results; **Middle panel**: Fully-adjusted results; **Right panel:** Betablocker only, subgroups by LVEF above or below 40% ACEi/ARB = angiotensin converting enzyme inhibitors or angiotensin receptor blockers; DAPT = dual antiplatelet therapy; LVEF= left-ventricular ejection fraction; OR = odds ratio

To better understand the relationship between PCa diagnosis and betablocker prescription, we looked at subgroup analysis by left ventricular ejection fraction (LVEF) as a post hoc analysis. We identified that the relationship was only statistically significant in those with an LVEF over 40% (OR 0.77; 0.67-0.90). In those with LVEF of 40% or less, there was no difference, however the confidence interval was wide.

## Discussion

### Summary of key findings

In this national, registry-based study we found no differences in the use of invasive management for AMI between patients with localized, non-metastatic PCa compared to the general AMI population. The use of secondary prevention after hospital discharge was also similar between patients with or without PCa. When looking at complications after treatment, we found a lower risk in the PCa group for composite complications, although a higher proportion of patients with PCa experienced serious bleeding during treatment for AMI. All-cause mortality up to one year after treatment was lower in patients with PCa, but there were no differences in MACE or re-infarction at 30 days or 1 year. There were no differences in MACE, re-infarction or mortality within the PCa population when comparing primary use of ADT.

### Interpretation

This study represents one of the first epidemiological studies comparing AMI treatment and outcomes in a nation-wide population with PCa to the whole general male, non-cancer AMI population with individual level data. This study benefited from high-quality registry data that utilized multiple sources of data to identify cancer cases, AMI cases, medical treatment, cause of death, comorbidity and sociodemographic characteristics for the entire male population over 40 years. This gave us the ability to control for important confounders, specifically prior CVD, smoking and cancer specific factors, reducing the risk of bias. We were also able to verify certain variables because we had data from multiple sources. For example, medications prior to the AMI, reported in NORMI, could be verified using data from NorPD. Results can be generalized to populations with similar demographics and access to health care.

Patients with localized PCa, whose average life expectancy is similar to that of the general population, should not receive different treatment for an AMI solely based on their cancer diagnosis ^(25)^. In contrast to our study findings, previous research has shown lower rates of invasive treatment in cancer patients, and worse outcomes following AMI, such as increased rates of in-hospital and long-term mortality ^(13–15)^. The primary difference in our study is that we have focused only on PCa and not mixed other types of cancers with different prognosis and CVD risk, which may explain our contradictory findings. However, differences in health care access and regional variations in the population, cannot be excluded as possible explanations for the differences.

The odds of death at 1 year and of experiencing an in-hospital complication was around 20% lower in the PCa population than in the general AMI population. We know that patients diagnosed with localized low- and intermediate-risk prostate cancer have a 5-and 10-year relative survival above 100%, implying that many people diagnosed with PCa are healthier than the age-matched background population ^(25, 26)^.

Norway does not currently have a national screening program for PCa but does benefit from universal health coverage. However, universal access to healthcare does not necessarily translate to uniform utilization rates across socioeconomic groups ^(27)^. A substantial number of patients with localized disease are detected through PSA testing in asymptomatic males and a higher frequency of PSA testing is observed among individuals belonging to higher socioeconomic classes, possibly attributed to improved healthcare system accessibility or varied utilization patterns ^(28)^.

In our study, one in five of the PCa patients that experienced an AMI were current smokers, compared to one in three in the general AMI population. Less comorbidities were also observed in the PCa patients in comparable age groups (**Supplemental 1**). Thus, it is likely that better general health among patients diagnosed with PCa may have influenced the risk of complications and mortality. Further, positive lifestyle changes after their PCa diagnosis, and treatment of modifiable CVD risk factors may also be of importance. Although we have adjusted for important confounders such as socioeconomic factors, comorbidity index and smoking, there might be differences between the populations that are not apprehended by our data.

Although overall complications were lower in PCa patients, 2.1% (30 patients) in the PCa group vs. 1.1% (375 patients) in the AMI group experienced serious bleeding during their hospital stay, corresponding to an estimated 60% increased risk for serious bleeding after AMI in adjusted analyses. Other studies, with mixed cancer types, have also found increased risk of bleeding in cancer patients after AMI invasive management ^(29)^. Cancer patients may specifically be at increased risk of bleeding for several reasons, such as increased use of prophylactic anticoagulants for their higher risk of venous thromboembolism (VTE), which is estimated to be nine times more prevalent in the cancer population compared to the general population ^(30)^. Furthermore, managing anticoagulant therapy for a VTE episode in individuals with solid tumors is challenging due to the heightened susceptibility to both thrombotic relapses and bleeding while undergoing treatment with anticoagulants ^(31, 32)^. In one study, major bleeding was one of the most common causes of death after VTE in cancer patients ^(33)^.

Cancer treatment, such as chemotherapy and radiotherapy, can increase the risk of bleeding ^(6)^. However, unlike other cancers, primary, localized PCa is not generally treated with chemotherapy, which can lead to reduced platelet count and thrombocytopenia ^(23)^. Radiotherapy can induce weakening of mucosal barriers and, specific for pelvic area radiotherapy, may result in damage to the surrounding bowels, increasing the risk of bleeding, potentially long-term ^(34)^. In our study population, using descriptive analyses, we did not find any higher rates of anticoagulation use prior to AMI in our PCa population or increased rates of bleeding in patients that received radiotherapy that would directly explain the in-hospital bleed. Further studies are needed to better understand the nature of the increased risk of bleeding in PCa patients.

Increased risk of bleeding complicates treatment of AMI. Common invasive management of AMI includes treatment with PCI, which requires long-term use of antiplatelet medications to reduce risk of thrombosis and occlusion at the stent, but also increases the risk of bleeding ^(29)^. Having information on the cancer patient’s risk of bleeding, such as history of VTE, use of anticoagulants, treatment with radiotherapy or chemotherapy and metastatic development, could help cardiologists determine appropriate treatment and use of DAPT. Results from recent studies have shown that in patients with high risk of bleeding and high risk of CVD, DAPT time can be reduced with extended monotherapy without a reduction in benefit ^(35, 36)^.

There is a global trend in reduced use of betablockers as secondary prevention after an AMI as more evidence implies that it does not benefit all patients. Findings from the REDUCE-AMI trial demonstrated that long-term betablocker use in patients with preserved ejection fraction did not improve all-cause mortality or CVD outcomes with a median 3.5 years of follow-up ^(37)^. Our analyses identified reduced prescription of betablockers in the PCa population, but after ad hoc analysis by left ventricular ejection fraction, the reduced betablocker use was only in those with preserved ejection fraction, where medical evidence for benefit is lacking.

### Study limitations

For PCa patients, we had detailed clinical and routine data during the diagnosis period, and limited information on the development of metastatic disease. We therefore focused on primary treatment, so there could have been ADT treatment or chemotherapy used for advancing disease, that we did not account for. We had limited clinical data to determine medical indication for AMI treatment (primary and secondary prevention), but any bias would likely be non-differential. To include as many PCa patients with AMI as possible and have sufficient power for our analyses, we included a long-time frame for cancer diagnosis (2004–2019). During this time there have been changes in treatment and diagnosis of PCa that could impact our findings. Use of new GnRH antagonists has become more prevalent globally, particularly in response to their being associated with limiting CVD risk. However, the use of these medications in Norway only began in the last few years and therefore the proportion in our study was extremely limited, with much less follow-up time. Therefore, we were not able to perform any specific analyses comparing GnRH agonists and antagonists.

## Conclusion

In our study comparing treatment and health outcomes after an AMI between PCa patients and the general AMI population, we identified no differences in invasive management or secondary medical treatment. While PCa patients had less risk of overall complications, they did experience increased risk of serious bleeding. PCa patients had better survival at 1 year, but no difference in MACE or re-infarction outcomes. There was no increased risk of MACE, re-infarction or mortality in PCa patients that received primary ADT treatment compared to no ADT treatment.

We conclude that there is no evidence for reduced quality of health care regarding AMI patients with PCa in Norway and support the recent updated ESC guidelines that recommend treatment of AMI as usual for PCa patients with a survival expectancy greater than 6 months, although with care to their increased risk of bleeding.

## Acknowledgements

Statistical analysis code in R was partly developed with the support of GPT UiO for trouble shooting and generating ideas. GPT UiO is a service developed by University of Oslo to use OpenAI’s GPT models within the university’s privacy requirements.

## Data availability

The data underlying this article cannot be shared publicly because the linkages were performed and delivered under a data permit that does not allow publicly sharing of data for the privacy of individuals included. The datasets can be identified and applied for through https://helsedata.no or can be shared upon reasonable request to the corresponding author, and after appropriate ethical approval have been granted.

